# The rise and global spread of IMP carbapenemases (1996–2023): a genomic epidemiology study

**DOI:** 10.1101/2025.05.25.25328332

**Authors:** Ben Vezina, Bhargava Reddy Morampalli, Hoai-An Nguyen, Angela Gomez-Simmonds, Anton Y. Peleg, Nenad Macesic

**Affiliations:** Department of Infectious Diseases, The Alfred Hospital and School of Translational Medicine, Monash University, Melbourne, Australia; Division of Infectious Diseases, Department of Internal Medicine, UC Davis Health, Sacramento, California, USA; Centre to Impact AMR, Monash University, Clayton, Australia; Infection Program, Monash Biomedicine Discovery Institute, Department of Microbiology, Monash University, Clayton, Australia; Infection Prevention & Healthcare Epidemiology, Alfred Health, Melbourne, Australia

## Abstract

**Background:** IMP carbapenemases confer extensive drug resistance and are increasingly noted worldwide. Despite this, little is known regarding the global epidemiology of IMP carbapenemases.

**Methods:** We comprehensively identified *bla*_IMP_ genes in all publicly available bacterial genomes, then systematically analysed the distribution of variants across species, lineages, plasmids and mobile elements, examining patterns over time, across geographic regions and by source. Structural analysis of IMP variants was performed.

**Findings:** 4,556 *bla*_IMP_-containing genomes were identified from 1996-2023, including 52 *bla*_IMP_ variants across 93 bacterial species. Key variants (*bla*_IMP-1_, *bla*_IMP-4_, *bla*_IMP-7_, *bla*_IMP-8_ and *bla*_IMP-13_) achieved global endemicity, while *bla*_IMP-26_ and *bla*_IMP-27_ were regionally endemic in Southeast Asia and North America, respectively. *bla*_IMP_ dissemination was driven by horizontal gene transfer, facilitating inter-species spread. Proliferation of multidrug-resistant *Enterobacter hormaechei*, *Pseudomonas aeruginosa* and *Klebsiella pneumoniae* lineages led to local outbreaks. Dereplication removed 3,175/4,556 (69.9%) genomes, indicating that most *bla*_IMP_-containing genomes were highly related. *bla*_IMP_ variants were associated with mobile genetic element combinations including class 1 integrons and insertion sequences (99.7%), aiding mobilisation into ≥52 plasmid clusters, predominantly IncHI2A, IncN, IncL/M and IncC. Genomes of environmental and animal origin accounted for 10.0% and 1.1% of the dataset, respectively. Evidence of cross-source transmission was limited, with most spillover occurring between genomes of human and environmental origin. Structural analysis revealed a conserved carbapenemase structure (mean lDDT 0.977), with convergent missense mutations at seven catalytically relevant sites.

**Interpretation:** Global analysis enabled us to historically reconstruct the emergence and variant-specific epidemiologies of *bla*_IMP_ carbapenemase genes. Intersecting mobile elements enabled *bla*_IMP_ genes to spread across multiple plasmids and bacterial genera, facilitating global and multi-source spread within a One Health framework. Additionally, convergent evolutionary patterns indicate that IMP variants may continue evolving, potentially evading novel beta-lactam antimicrobial agents.

**Funding:** NHMRC EL1 (APP1176324) to N.M.; NHMRC PF (APP1117940) to A.Y.P.; NIH/NIAID R01AI175414 to A.G-S.

**Research in context panel:** *Evidence before this study:* Despite being a major cause of carbapenem resistance in Gram negative infections, little is known about the global epidemiology of IMP carbapenemases. IMP carbapenemases are metallo-beta-lactamases that were first identified in 1991 and have evolved into 96 different IMP variants. On May 21 2025, we searched all published reports available in PubMed using the terms “’IMP’ and ‘carbapenemase’ genomics NOT (Review[Publication Type]) NOT (Case Reports[Publication Type]) NOT PCR” with no language restrictions and no publication date restrictions. We identified 223 articles, 62 and 121 of which reported single species or a single study centre/country, respectively. Only 6 articles employed genomics to examine multi-species and multi-geographical isolates, though this was in the context of carbapenem resistance more broadly rather than IMP carbapenemases specifically. The most relevant study included 38 globally distributed genomes across four species and tracked seven blaIMP variants across mobile genetic elements.

*Added value of this study:* To our knowledge, this global characterisation provides the most comprehensive account of *bla*_IMP_ carbapenemase gene epidemiology. To analyse the global distribution and diversity of *bla*_IMP_ genes, we compiled all available public genome data resulting in a dataset of 4,646 genomes. This has allowed us to identify local, regional and international spread of *bla*_IMP_ variants and determine the contributions of clonal expansion, plasmid proliferation and co-localised mobile genetic elements. We demonstrated that key *bla*_IMP_ variants display global (IMP-1, IMP-4, IMP-7, IMP-8 and IMP-13) and regional (IMP-26 within Southeast Asia and IMP-27 within North America) endemicity and that these patterns have been previously unacknowledged, reframing the previous understanding that IMP carbapenemases were largely confined to the Asia-Pacific region. Our observation of convergent evolutionary patterns raise concern that IMP variants may continue to evolve, potentially evading new β-lactam antimicrobials. This analysis has revealed the under-recognised contribution IMP carbapenemases make to global carbapenem resistance.

*Implications of all the available evidence:* These findings provide the first comprehensive atlas of *bla*_IMP_ carbapenemase gene dissemination and underscore the silent global spread of IMP carbapenemases. We note the critical need for enhanced surveillance systems, particularly in low- and middle-income countries, that can detect complex plasmid-mediated and mobile genetic element-associated spread, as we noted with *bla*_IMP_ carbapenemase genes. Moreover, our analyses show that systematic sampling across human, animal, and environmental reservoirs is crucial to address the One Health dimensions of emerging antimicrobial resistance threats. The study provides a framework for future interventions aimed at tracking and stopping the spread of IMP carbapenemases and calls for co-ordinated, real-time public health responses to this growing challenge.

## Introduction

Carbapenemase-producing organisms (CPOs) are a significant threat to global health and have been deemed critical priority pathogens by the World Health Organization (1). Five key carbapenemase classes (KPC, NDM, OXA, VIM and IMP) cause the majority of global infections (2,3). Despite being included amongst these, little is known about the global epidemiology of IMP carbapenemases. IMP carbapenemases are metallo-beta-lactamases (MBLs) that were first identified in 1991 in *Pseudomonas aeruginosa* in Japan (4). To date, 96 different IMP carbapenemase variants have been identified and IMP carbapenemases are now endemic to Asia and Australia (5–10). In addition, outbreaks of IMP-carrying organisms are increasingly reported across several regions including Europe and the Americas (11–15). This is a highly concerning development given the paucity of treatment options for infections caused by these organisms, including resistance to novel agents with activity against other MBLs such as cefepime-taniborbactam (16).

Carbapenemases disseminate through various mechanisms, including transposon-mediated transfer between plasmids (*bla*_NDM_), stable association with successful clonal lineages (*bla*_KPC_), rapid expansion of a single epidemic plasmid across multiple bacterial lineages (*bla*_OXA-48_), and transient associations involving diverse plasmids and numerous lineages (17,18). Our prior work indicated that in Australia *bla*_IMP-4_ spreads both clonally and through horizontal transfer via mobile genetic elements (19). However, current data on *bla*_IMP_ dissemination remain limited: most prior studies have focused on a single IMP carbapenemase type and/or a specific geographical region (5,19–23).

We therefore aimed to comprehensively determine the genomic epidemiology of *bla*_IMP_ carbapenemase genes. Specifically, we dissected the dynamics underlying *bla*_IMP_ dissemination and evaluated the contributions of genomic factors, outbreak events, structural determinants and One Health-related influences. We analysed all publicly-available *bla*_IMP_-carrying genomes (n=4,556) spanning almost three decades (1996–2023), uncovering global expansion and regional endemicity of diverse *bla*_IMP_ variants. Collectively, this work creates an atlas of *bla*_IMP_ carbapenemase genes that highlights their transition from initial endemic foci in the Asia-Pacific region to a worldwide public health threat and emphasises the pressing need for integrated strategies to combat their further spread.

## Results

We identified 4,556 genomes (4,020 assembled from short- and 536 from long-read sequencing data) isolated globally from 1996-2023 carrying 52 distinct *bla*_IMP_ variants across 26 bacterial genera (**Fig. 1** and **Table S1**). This revealed a remarkable diversity of both *bla*_IMP_ genes and their bacterial hosts, totalling 4,559 total *bla*_IMP_ genes, with three genomes carrying two *bla*_IMP_ variants each. *bla*_IMP-4_ and *bla*_IMP-1_ were the most frequent variants, found in 1,592/4,559 (34.9%) and 1,155/4,559 (25.3%) genomes, respectively (**Table S2**). The most prevalent species included 1,053 *Enterobacter hormaechei* (23.1%), 977 *Pseudomonas aeruginosa* (21.4%) and 681 *Klebsiella pneumoniae* (14.8%), together accounting for 59.4% of the dataset. Of the 4,556 *bla*_IMP_-carrying genomes, 728 (16%) carried mobile colistin resistance (*mcr*) genes and 345 (7.6%) carried other carbapenemase genes (**Table S1**).

**Fig. 1:**
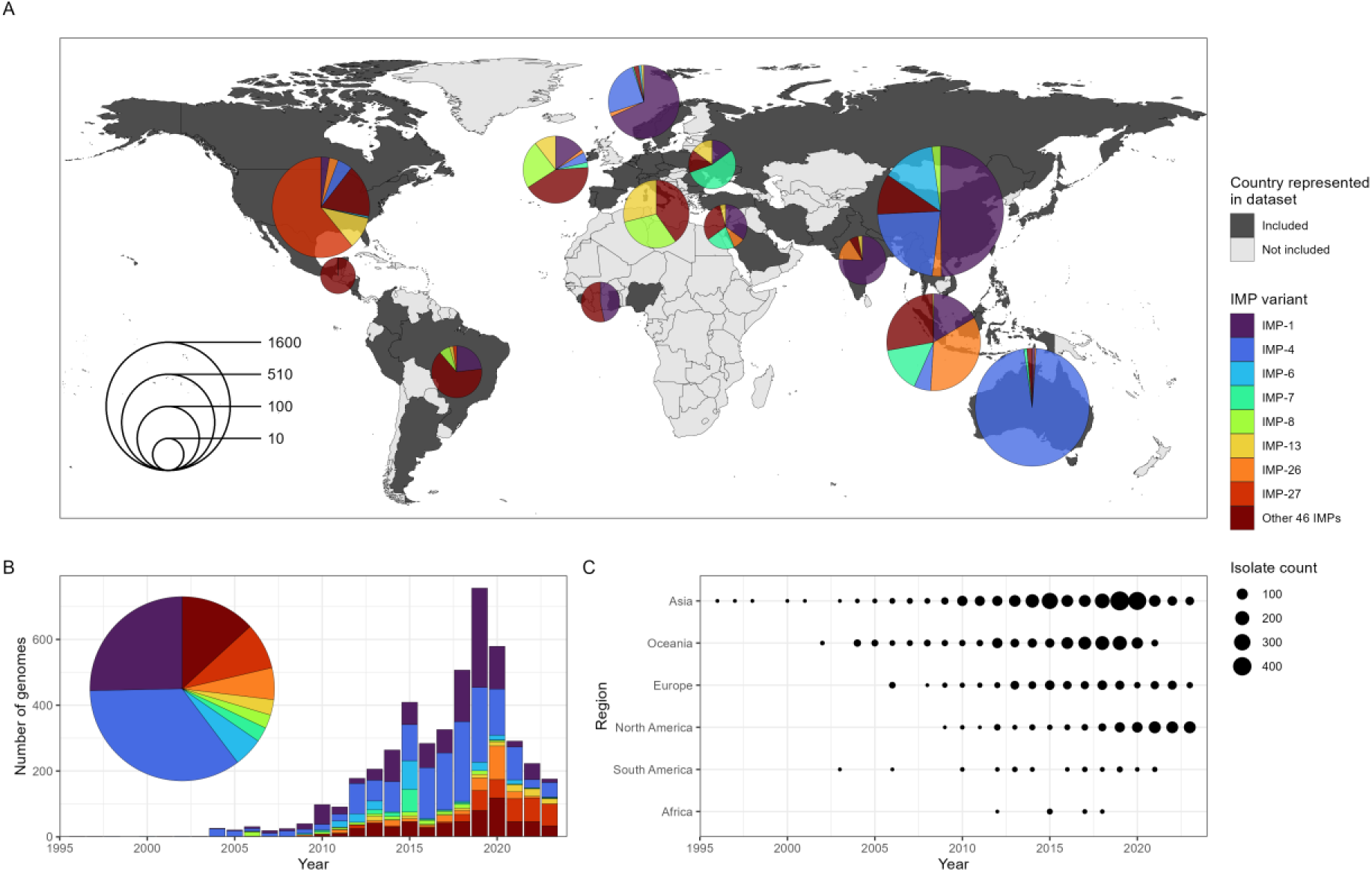
Geographical and temporal spread of *bla*_IMP_ carbapenemase genes. **A:** World map showing breakdown of *bla*_IMP_-carrying genomes and their global species distribution. Size of pie charts indicates number of isolates. *bla*_IMP_ variants with <100 total observations are grouped into ‘Other 46 IMPs’. **B:** Global prevalence of most common *bla*_IMP_ variants over time. **C:** Dot plot showing prevalence of *bla*_IMP_-positive genomes within the United Nations geoscheme regions over time.

### Tracking the global spread of *bla*IMP carbapenemase genes

Our dataset enabled us to reconstruct the global expansion of *bla*_IMP_ genes from origins in the Asia-Pacific region to an increasing number of variants detected across multiple regions as defined by the United Nations geoscheme (**Fig. 1A, C**).

From 1996-2011, *bla*_IMP-1_, *bla*_IMP-4_ and *bla*_IMP-6_ emerged in Asia and Australia, together accounting for 79.44% genomes studied. *bla*_IMP-1_ was identified across Eastern and South Eastern Asia (939/1,155 *bla*_IMP-1_ genomes) during the entire study period, most predominantly Japan, Singapore and China. After 2014, regional *bla*_IMP-1_ outbreaks were increasingly noted outside of Asia, including in Europe, Western Africa and North America. *bla*_IMP-4_ was initially noted in China in 1998 and continued to be isolated there through the study period (379/1,592 *bla*_IMP-4_ genomes). However, from 2002 it was well established in Australia (1,105/1,592 genomes), predominantly on the east coast. Similarly to *bla*_IMP-1_, sporadic *bla*_IMP-4_ outbreaks were noted outside these regions between 2014-2023 in Europe and Northern America (62/1,592 *bla*_IMP-4_ genomes). *bla*_IMP-6_ was identified in Japan from 2000 and remained focused there until end of the study period (224/235 *bla*_IMP-6_ genomes), with sporadic isolation in South Korea, UK and USA between 2017-2023 (n=11 genomes).

From 2009, *bla*_IMP-26_ and *bla*_IMP-27_ emerged as regional *bla*_IMP_ genes in South Eastern Asia and North America, respectively (**Fig. 2**). *bla*_IMP-26_ was first noted in 2009 and became established in *P. aeruginosa* in Vietnam and Philippines. *bla*_IMP-26_ was highly associated with *P. aeruginosa* ST235 (151/254 *bla*_IMP-26_ genomes). This variant-lineage combination led to subsequent spread internationally across six geographic subregions from South-Eastern Asia to Southern Asia, Australia, Northern Europe, Western Europe and Northern America from 2009-2022, possibly due to travel-associated importation (**Fig. 2A**, **Table S3**). *bla*_IMP-27_ emerged from 2011 as the dominant carbapenemase in North America, specifically the US (genomes from 21 states) (**Fig. 2B**). In addition to geographical location, this *bla*_IMP_ variant displayed a distinct epidemiology characterised by associations with agricultural animals from 2011-2014 before emerging in human-origin genomes from 2016 onwards (**Table S3**). 77.6% (287/370 North American *bla*_IMP-27_ genomes resulted from expansion of local lineages of *Providencia rettgeri* spp. 1, 2 and 3, *Providencia stuartii* spp. 1 and 2, *Providencia huaxiensis*, *Proteus mirabilis* and *Morganella morganii*.

**Fig. 2:**
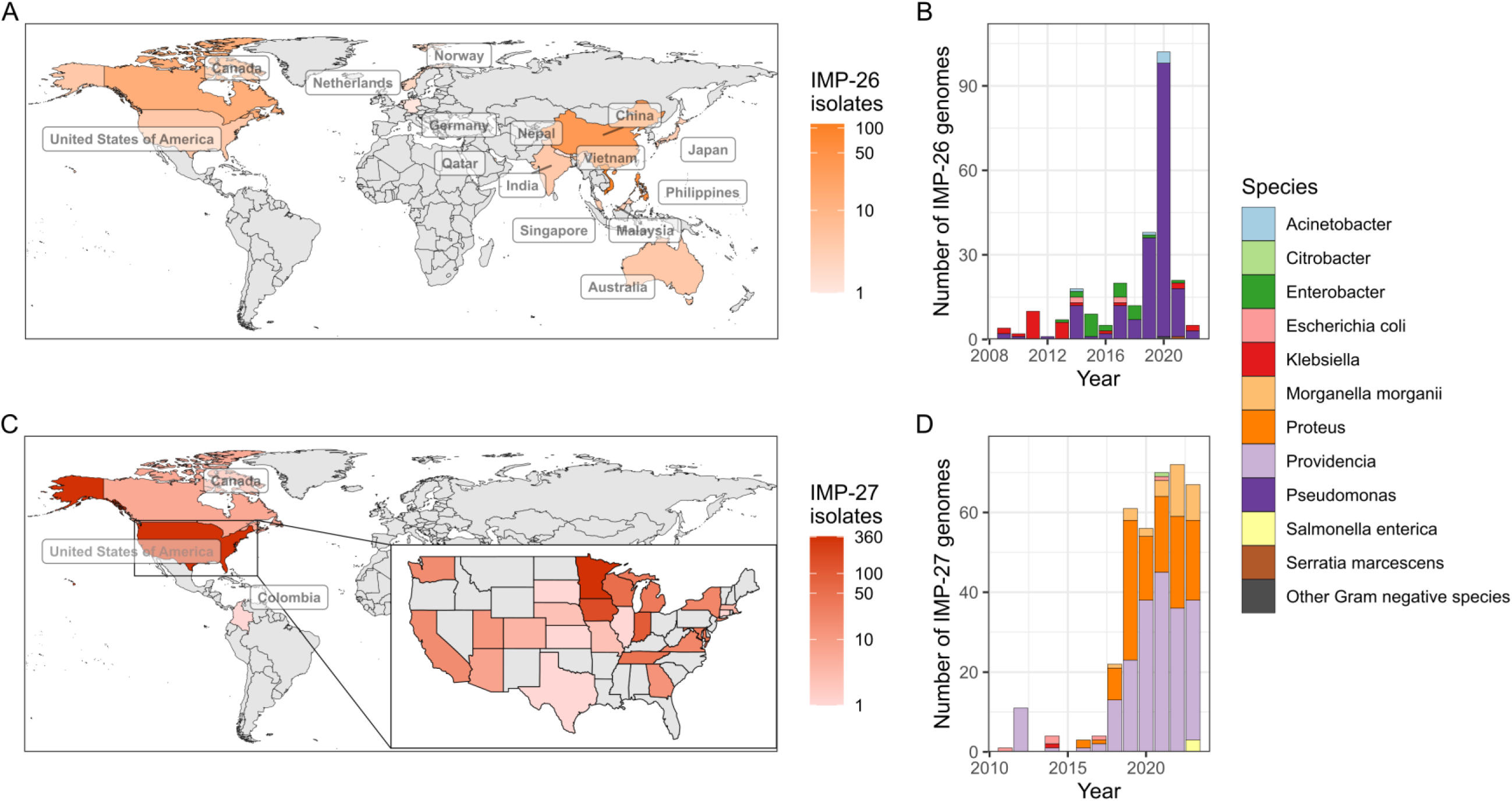
*bla*_IMP-26_ and *bla*_IMP-27_ epidemiology. **A:** Global distribution of *bla*_IMP-26_, showing regional endemicity in Asia and putative travel-association with other geographic regions. **B:** Species breakdown of *bla*_IMP-26_ genomes over time. **C:** Global distribution of *bla*_IMP-26_, with panel showing regional endemicity concentrated within the US. **D:** Species breakdown of *bla*_IMP-27_ genomes over time.

From 2018 to 2023, several *bla*_IMP_ variants achieved global (*bla*_IMP-1_, *bla*_IMP-4_, *bla*_IMP-7_, *bla*_IMP-8_, *bla*_IMP-13_) and regional (*bla*_IMP-6_, *bla*_IMP-26_, *bla*_IMP-27_) endemicity, whereby there was evidence of ongoing spread outside of sporadic outbreaks (**Table S4**). During this period, the global dissemination of *bla*_IMP_ carbapenemase genes was demonstrated with their detection in 42 countries spanning all regions, with 23 countries reporting ≥2 *bla*_IMP_ variants and 8 countries reporting ≥5 *bla*_IMP_ variants.

### *bla*IMP carbapenemase genes found in diverse bacterial hosts with over-representation of multidrug-resistant lineages

Having determined that *bla*_IMP_ genes achieved global spread, we wanted to understand how genomic factors shaped this spread and hence adopted a multi-level approach focusing on bacterial hosts, plasmids and finally other mobile genetic elements. On a bacterial host level, there were 93 species carrying 4,556 total *bla*_IMP_ genes but we noted that *bla*_IMP_ variants were associated with specific species (**Fig. 3A, Table S5**). *bla*_IMP-1_, *bla*_IMP-4_ and *bla*_IMP-6_ were predominantly associated with *Enterobacterales* (specifically *E. hormaechei*, *K. pneumoniae, E. coli*), while *bla*_IMP-7_, *bla*_IMP-13_ and *bla*_IMP-26_ were predominantly noted in *P. aeruginosa*. *bla*_IMP-27_ had a unique epidemiology dominated by *Providencia, Proteus* and *Morganella* spp.

**Fig. 3:**
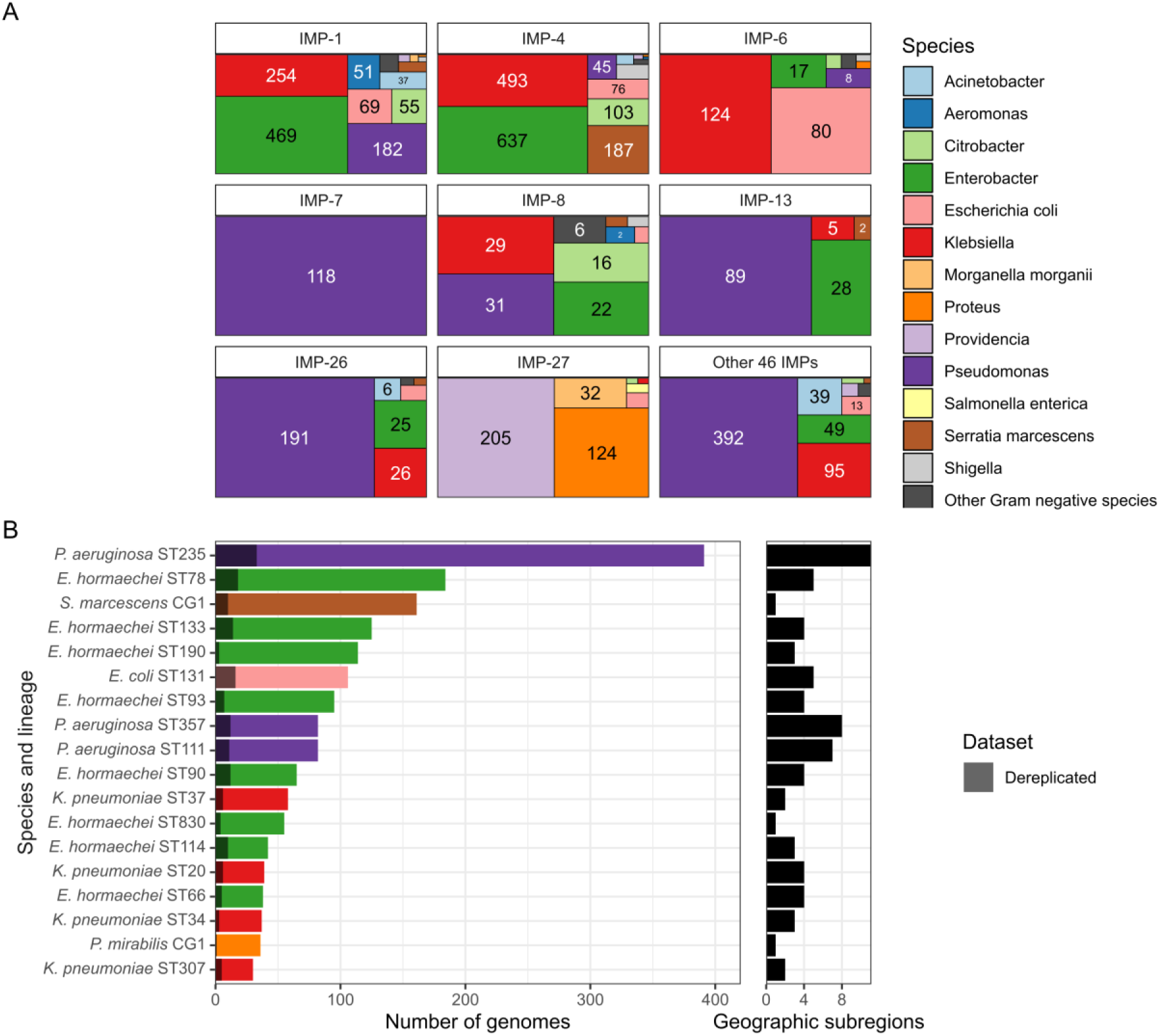
Species and lineages associated with *bla*_IMP_ carriage. **A:** Species breakdown across each *bla*_IMP_ variant. **B:** Highly prevalent lineages within the dataset, showing the impact of dereplicating IMP-clusters. Dereplicated bars are shown as grey overlays over coloured genome counts, not stacked bars. Only lineages with ≥30 genomes are shown. Number of geographic subregions each lineage was detected in is shown as a companion plot. Full data found in **Table S5** and **Table S6.**

**Fig. 4:**
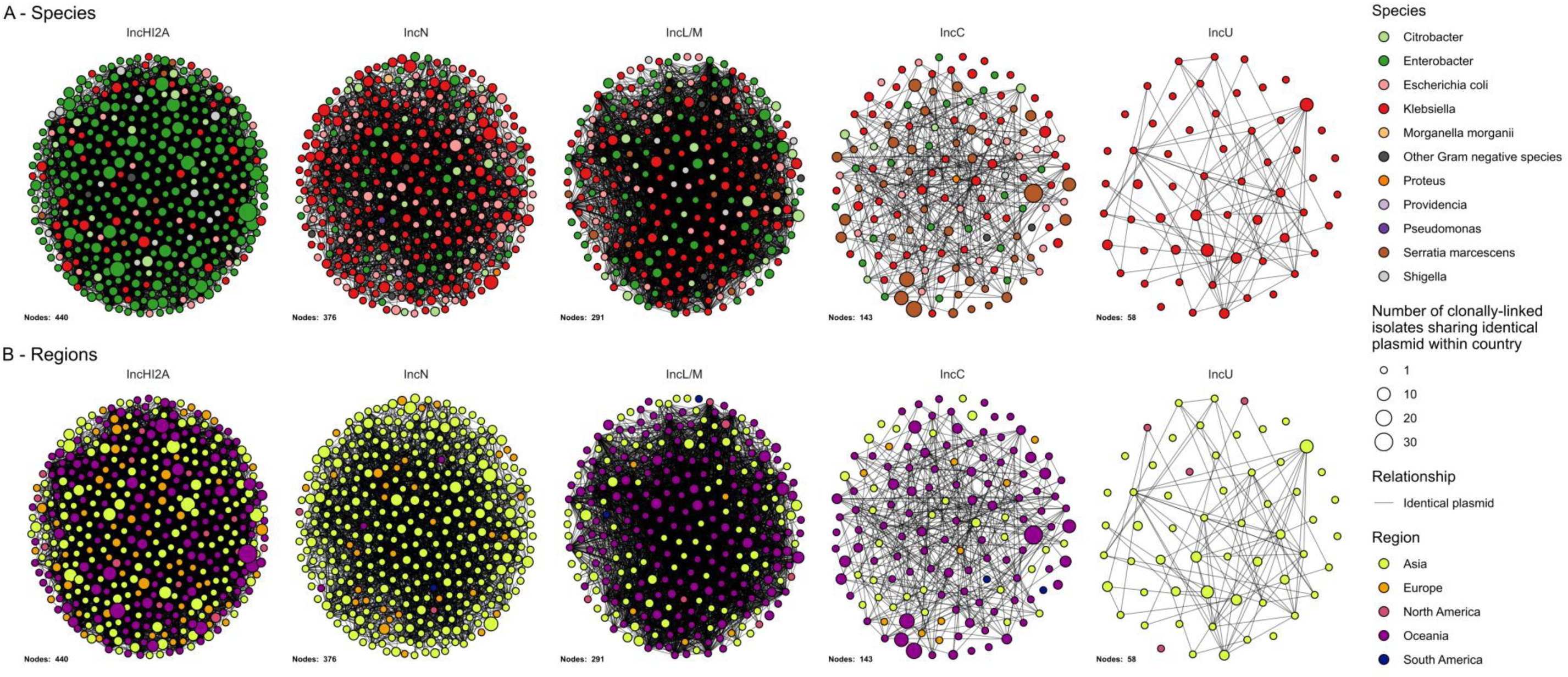
Key *bla*_IMP_ plasmids by bacterial species and geographic region. **A:** Network of related plasmids, separated by plasmid Inc types and coloured by host species. Edges show identical plasmids between IMP-clusters (nodes). Size of node represents number of genomes within an IMP-cluster and country also carrying the same plasmid. **B:** Identical networks but coloured by geographical region.

*P. aeruginosa* sequence type ST235 was the most frequent single lineage isolated (391/4,556 genomes, 8.6%) and carried the greatest number of *bla*_IMP_ variants (n=17) (**Table S5**). *P. aeruginosa* ST235 is a global multidrug-resistant (MDR) lineage, recognised for its ability to harbour a high diversity of acquired resistance genes (24). While there was a close association with *bla*_IMP-26_ (151/391 *P. aeruginosa* ST235 genomes), ST235 also carried other key *bla*_IMP_ variants including *bla*_IMP-1_ (55/391 genomes), *bla*_IMP-51_ (51/391 genomes), and *bla*_IMP-7_ (46/391 genomes). We noted *bla*_IMP_ presence in several other MDR lineages: *E. hormaechei* ST78 had a close association with *bla*_IMP-1_ (162/4,556 [3.6%] genomes, 160 from Japan); *E. coli* ST131 with *bla*_IMP-6_ (64/4,556 [1.4%] genomes, all from Japan); *K. pneumoniae* ST307 harboured *bla*_IMP-38_ in a limited number of genomes (n=22, 21/22 from China) (**Fig. 3B**, **Table S6**).

We then quantified the impact of clonal bias and potential outbreaks by only including a single representative from each cluster of closely-related genomes (i.e. ‘dereplication’) (see **Methods**). We defined these ‘IMP-clusters’ as clusters of genomes which shared the same *bla*_IMP_ variant, species, lineage (ST or clonal group [CG], if no MLST schema available) and were within a species-specific threshold of 5 single nucleotide variants (SNV) per Mb (see **Methods**). This resulted in 1,381 IMP-clusters, with 3,175/4,556 (69.9%) closely-related genomes removed by dereplication (**Fig. 3B**, **Table S6**). Indeed, the majority of *bla*_IMP_ genomes were clonally linked to at least one other genome: 700/1,381 IMP-clusters contained >1 genome (**Fig. 3B, Fig. S1**), potentially reflecting sequencing conducted in outbreak settings. This included multiple *P. aeruginosa* ST235 IMP-clusters (*bla*_IMP-26_ - 127 genomes, *bla*_IMP-51_ - 51 genomes, *bla*_IMP-1_ - 50 genomes, *bla*_IMP-31_ - 39 genomes) (**Fig. S2A, Table S6**). Some IMP-clusters also accounted for high proportions of observations for that gene: *K. pneumoniae* ST37 and *E. coli* ST131 *bla*_IMP-6_ IMP-clusters accounted for 14.5% and 13.2% of all *bla*_IMP-6_ observations, while *E. hormaechei* ST190 and ST78 IMP-clusters accounted for 6.91% and 6.67% of *bla*_IMP-4_ and *bla*_IMP-1_ observations. We then assessed IMP-clusters for possible spread between countries and geographic subregions and noted that only 108/1,381 (7.8%) were found across >1 country. From the perspective of regional spread, 5.7% (78/1,381) IMP-clusters were found across ≥2 geographic regions, including 24 independent *P. aeruginosa* IMP-clusters (**Table S6**).

### Diverse plasmids facilitate global dissemination of *bla*IMP carbapenemase genes

To determine the genetic context of *bla*_IMP_ genes, we first analysed all available long-read genomes in the dataset (n=539) (see **Methods**). *bla*_IMP_ genes were located on plasmids in 436/539 genomes (80.9%). These were divided into 52 plasmid ‘clusters’ as defined by MOB-suite plasmid replicon clusters (25), which carried a total of 20 *bla*_IMP_ variants (**Table S7**). IncHI2A, IncC and IncN plasmids collectively accounted for 56.0% (244/436) plasmids and each carried four, two and five different *bla*_IMP_ variants. These plasmids were found across Asia and Australia with sporadic cases in Europe (**Table 1** and **Table S7**). Other key *bla*_IMP_ plasmids were IncU, IncFIB, IncFIB/IncFII-type 2, IncL/M, IncP and an untyped *A. baumannii* plasmid. These carried 2-4 *bla*_IMP_ variants and were found in ≥2 geographic regions.

**Table 1:** Summary of key plasmid clusters (n≥100) and associated features. Full data can be found in Table S1 and Table S7.

| MOB-suite cluster | Inc replicon | Year range | *bla*<sub>IMP</sub> variant | Countries | Species |
| --- | --- | --- | --- | --- | --- |
| AA739_AJ055 | IncHI2A (n=396) | 2004-2022 | IMP-4, IMP-26, IMP-1, IMP-22, IMP-13 | Australia, Philippines, Japan, Ireland, UK, Spain, US | *K. michiganensis*, *K. oxytoca*, *E. hormaechei*, *K. pneumoniae*, *E. cloacae*, *C. youngae*, *E. asburiae*, *E. kobei*, *R. planticola*, *E. roggenkampii*, *E. coli*, *C. amalonaticus*, *K. aerogenes*, *C. freundii*, *E. bugandensis*, *K. variicola*, *S. boydii*, *S. sonnei*, *C. koseri*, *C. farmeri*, *E. chengduensis*, *C. portucalensis* |
| AA552_AI753 | IncN (n=302) | 2004-2023 | IMP-6, IMP-1, IMP-66, IMP-26, IMP-22, IMP-4, IMP-74, IMP-11 | Japan, Philippines, UK, Singapore, Spain, Portugal, Germany, Peru, China | *K. pneumoniae*, *E. coli*, *C. freundii*, *E. asburiae*, *E. hormaechei*, *E. kobei*, *E. ludwigii*, *K. michiganensis*, *R. ornithinolytica*, *K. variicola*, *S. sonnei*, *E. chengduensis*, *P. stuartii* Spp 1, *L. adecarboxylata*, *P. mirabilis*, *K. quasipneumoniae*, *E. cloacae*, *K. grimontii*, *C. braakii* |
| AA739_AJ057 | IncHI2A (n=276) | 2007-2021 | IMP-1, IMP-13 | Japan, US, UK | *E. hormaechei*, *C. freundii*, *E. asburiae*, *E. kobei*, *K. michiganensis*, *E. cloacae*, *E. coli*, *E. chengduensis*, *K. pneumoniae*, *S. marcescens*, *E. roggenkampii*, *K. oxytoca* |
| AA860_AJ266 | IncC (n=260) | 2002-2022 | IMP-4, IMP-1, IMP-60, IMP-8, IMP-15, IMP-23 | Australia, Japan, Netherlands, UK, Spain, China, Brazil, Singapore | *S. marcescens*, *K. pneumoniae*, *E. chengduensis*, *E. hormaechei*, *K. oxytoca*, *C. freundii*, *E. asburiae*, *E. coli*, *V. alginolyticus*, *K. michiganensis*, *K. variicola*, *S. sonnei*, *E. cloacae*, *P. mirabilis*, *C. koseri*, *E. bugandensis*, *K. quasipneumoniae* |
| AA002_AH532 | IncL/M (n=253) | 2006-2023 | IMP-4, IMP-34, IMP-1, IMP-22, IMP-59 | Australia, Japan, Philippines, Portugal, US, UK | *K. pasteurii*, *K. quasipneumoniae*, *C. freundii*, *K. pneumoniae*, *E. hormaechei*, *E. coli*, *K. michiganensis*, *S. sonnei*, *S. marcescens*, *C. murlinae*, *K. aerogenes*, *E. asburiae*, *K. variicola*, *S. boydii*, *C. koseri*, *E. kobei*, *L. adecarboxylata*, *A. subterranea*, *C. farmeri* |
| AA552_AI757 | IncN (n=153) | 2008-2023 | IMP-6, IMP-4, IMP-38, IMP-26 | Japan, China, US | *K. pneumoniae*, *K. michiganensis*, *K. pasteurii*, *A. subterranea*, *C. freundii*, *E. asburiae*, *E. hormaechei*, *K. grimontii*, *E. coli*, *E. soli*, *R. ornithinolytica*, *K. quasipneumoniae*, *K. variicola*, *R. planticola*, *C. amalonaticus*, *C. portucalensis*, *H. chinensis*, *C. braakii*, *E. roggenkampii* |
| AA739_AJ059 | IncHI2A (n=134) | 2008-2023 | IMP-6, IMP-1, IMP-4, IMP-22, IMP-8, IMP-13, IMP-26, IMP-19 | UK, Portugal, Taiwan, US, China, Australia, Poland | *C. youngae*, *E. hormaechei*, *E. asburiae*, *A. hermannii*, *E. cloacae*, *E. coli*, *K. pneumoniae*, *E. bugandensis*, *K. oxytoca*, *C. freundii*, *E. kobei*, *K. aerogenes*, *S. marcescens*, *P. vulneris* |
| AA739_AJ058 | IncHI2A (n=113) | 2010-2022 | IMP-4, IMP-6, IMP-1, IMP-13 | Australia, Japan, Spain, US, Singapore, China | *E. hormaechei*, *K. oxytoca*, *K. pneumoniae*, *E. coli*, *E. asburiae*, *K. michiganensis*, *E. kobei*, *C. amalonaticus*, *K. variicola* |

There were 181 plasmid cluster-*bla*_IMP_ variant combinations and we evaluated their geographic distributions. We noted diversity of IncN, IncHI2A and IncP plasmids, with multiple distinct plasmid clusters (n=8, n=5 and n=4, respectively). These were predominantly associated with one *bla*_IMP_ variant and one region (**Table S7**). Only 54/181 *bla*_IMP_ variant-plasmid cluster combinations were found across different countries, while 13/181 were found in ≥3 geographic regions, driven by successful expansion of key IMP-clusters carrying broad host-range plasmids (IncHI2A, IncL/M, IncC, IncN). This indicated that while subregional spread of closely-related plasmids may have occurred, spread between regions was not detected in most cases. The regional *bla*_IMP-26_ gene was found in IncHI2A, IncU and five untypeable plasmids (MOB-suite clusters AA739_AJ059, AC212_AL309, AC213_AL312. AC935_AM305 and AD068_AM495), respectively, however long read data was limited (13 genomes). Plasmids also displayed clear, species-specific host ranges. Most notably, IncHI2A plasmids were overwhelmingly associated with *E. hormaechei* (n=101), while IncC plasmids were associated with *S. marcescens* (n=30) and *K. pneumoniae* (n=18).

To gain insights from genomes with short-read data only, we clustered short-read contigs to long-read plasmid clusters (see **Methods**), with the caveat of reduced predictive confidence. In this analysis, *bla*_IMP_ variants were found on plasmids in 2,909/4,559 (63.8%) genomes, the chromosome in 97 genomes, with 1,553 remaining unclassified (**Table S7**). Representation of most plasmids was similar between following inclusion of short-read assemblies (**Fig. S3**). Notable exceptions included IncC plasmids decreasing from 16.3% to 8.3%, while IncN and IncL/M plasmids increased (10.8% to 19.7% and 8.3% to 14%, respectively). IncN increases resulted from short-read *K. pneumoniae* species complex genomes largely absent in the long-read/hybrid dataset, while the IncL/M increase was driven by additional *Enterobacter* spp., *K. pneumoniae* species complex and *E. coli* short-read genomes. Of the 181 *bla*_IMP_ variant-plasmid cluster combinations, 55 were found across multiple countries. A key example were *bla*_IMP-4_ IncL/M plasmids that were detected in 249 genomes across Australia, US, UK and Philippines in 19 bacterial species (predominantly *Enterobacterales*). The remaining 126/181 *bla*_IMP_ variant-plasmid cluster combinations were specific to a single country.

We then assessed relationships of *bla*_IMP_ plasmids with bacterial host lineages. In our prior work, we noted successful lineage-plasmid pairings that we termed ‘propagators’ (19). There were 35 lineages possibly acting as propagators with ≥10 genomes including *S. marcescens* CG1 carrying *bla*_IMP-4_ IncC plasmids (145 genomes), and *bla*_IMP-1_ IncHI2A plasmids associated with *E. hormaechei* ST78 and *E. hormaechei* ST133 (n=116 and n=88, respectively) (**Table S8**). Additionally, we identified 64 ‘connector’ lineages capable of harbouring ≥3 *bla*_IMP_ plasmid clusters (**Table S8**), which could serve as an opportunity for transfer of *bla*_IMP_-containing integrons (19). *E. hormaechei* ST78 and *E. coli* ST131 were the most prominent, carrying 7 plasmid clusters each (**Table S8**).

### Mobile elements associated with *bla*IMP carbapenemase genes

*bla*_IMP_ genes are frequently associated with integrons (26), integrative and conjugative elements (ICEs) (27), insertion sequences (IS) (28) and transposons (29). We therefore sought to determine if these mobile elements could provide insight into the spread of *bla*_IMP_ genes. We analysed the genetic context 10kb up- and downstream of each *bla*_IMP_ gene. To maximise identification of mobile elements, only contigs ≥6kb in length were considered for this analysis which resulted in 2,314/4,559 eligible *bla*_IMP_-containing contigs (**Fig. 5**). Initially, we examined the total presence of mobile elements across the dataset. Of these, 2,053/2,314 (88.7%) contigs showed direct association with integron elements (850 intact and 1,203 with *attC* clusters only) and almost all were class 1 integrons (795/850). When taking other mobile elements into account, integrons were found alone in 478/2,314 (20.6%) contigs but were co-located with other mobile elements in the majority, including IS (1,332/2,314 [57.6%]) and ICE (192/2,314 [8.3%]) (**Fig. 5**).

**Fig. 5:**
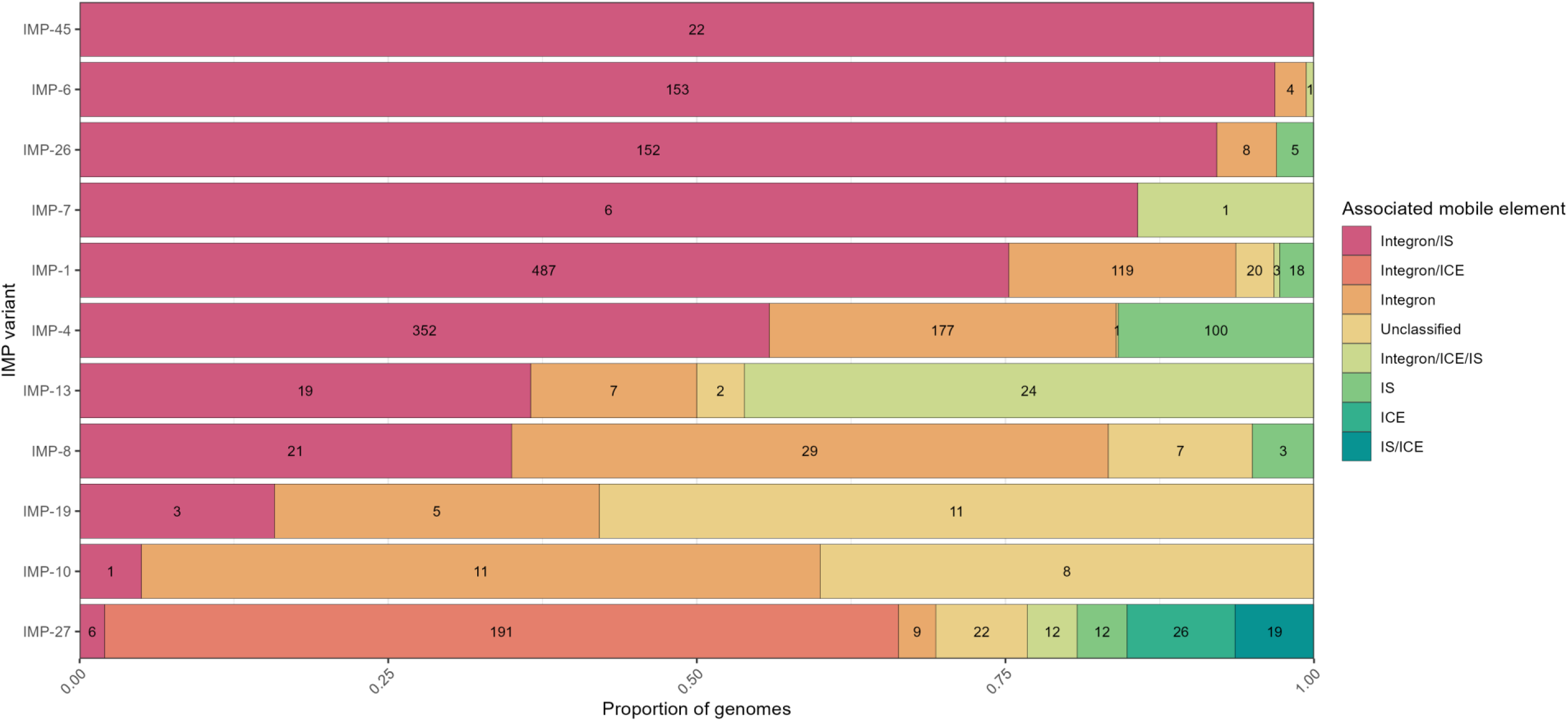
The majority of *bla*_IMP_ variants are associated with integrons and IS elements. Column graph showing the proportion of each *bla*_IMP_ variant and its association with mobile elements including integrative conjugative elements (ICE), integrons, insertion sequences (IS), and unclassified. Numbers within bars show *n* of each group. Full data found in **Table S1**.

Co-location of multiple mobile elements with *bla*_IMP_ genes may have contributed to their dissemination across populations due to increased mobility. Integron/IS-associated *bla*_IMP_ genes were most frequent (across 56 species, 39 IMP variants and 41 plasmid clusters), dating back to the earliest *bla*_IMP_ isolates in the late 1990s (**Fig. S4**). We saw no major differences between *bla*_IMP_ variants and their putative associated mobile element/s. *bla*_IMP-27_ was a notable exception: it had the most variable mobile element associations, characterised by presence of ICE alone or in combination with integrons and/or other IS (**Fig. 5**).

### IMP variants display convergent evolutionary trends

Beyond analysing genomic contributors to IMP dissemination, we analysed sequence and structural variation of IMP variants in our study (**Fig. 6A and B**). Critically, this variation is closely linked with β-lactam hydrolytic specificity and alters minimum inhibitory concentrations of carbapenems (30–33). This led us to hypothesise that it may influence *bla*_IMP_ dissemination. We noted IMP sequences were highly structurally conserved, with a mean local distance difference test (lDDT) score of 0.977 (**Fig. 6C**). Despite this, we saw reduced amino acid conservation scores at key positions throughout the structure (**Fig. 6D**), indicative of varying substrate specificity/activity. We examined mutations in key residue positions (30, 31, 51, 87, 150, 167, 196) previously shown to impact carbapenem hydrolytic specificity and noted a consistent pattern of convergent evolutionary mutations throughout the protein phylogeny at these key residues. Many of these convergent mutations appear to have been acquired independently (**Fig. 6A**), such as 31F found in multiple IMP sequences.

**Fig. 6:**
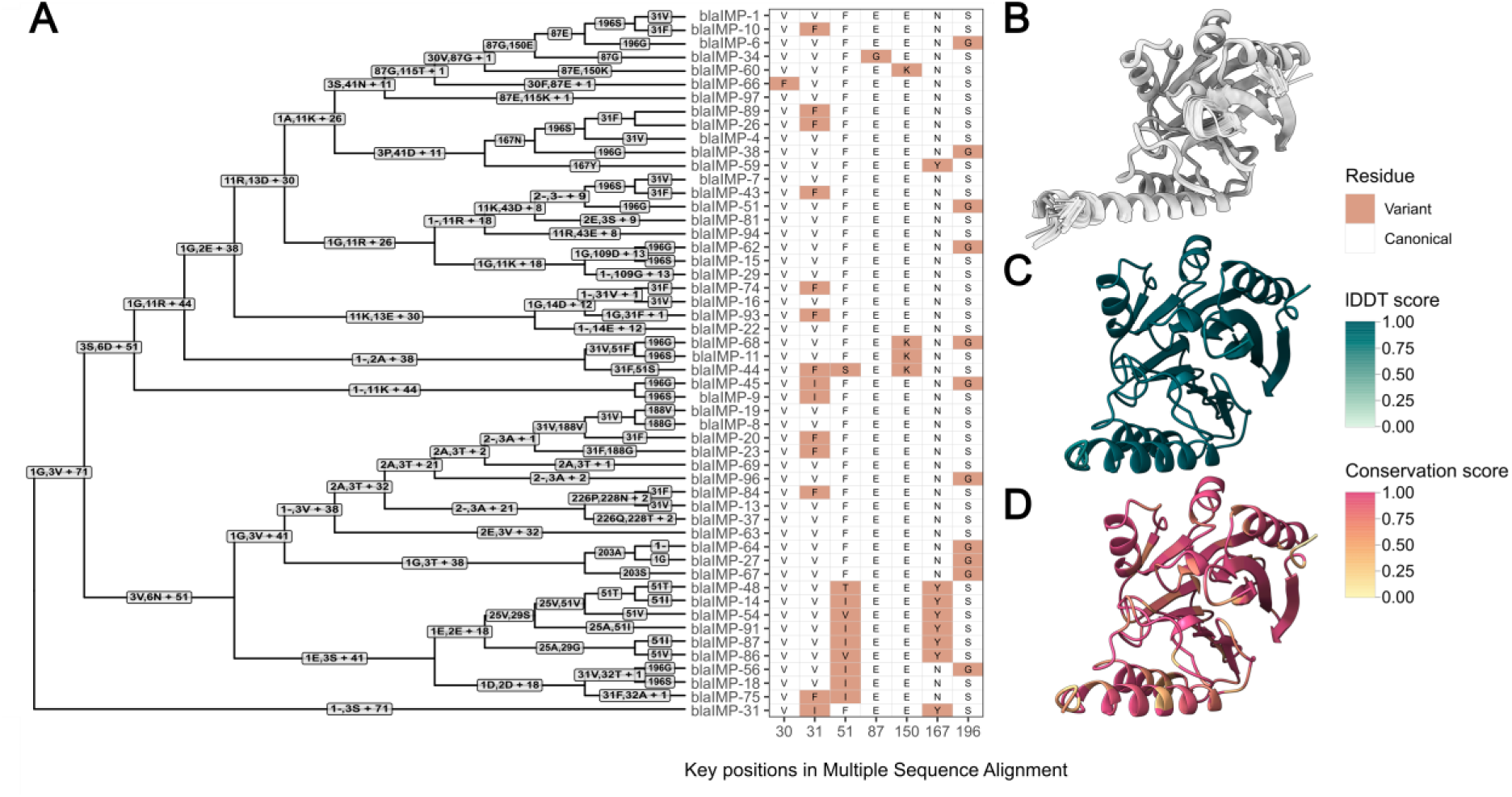
IMP variants have key residue changes which alter catalytic activity. **A:** Multiple sequence alignment guide tree with associated amino acid changes. Key residues which have demonstrated impact on antimicrobial resistance profiles (30,33–35) are shown as a heatmap. **B:** Predicted AlphaFold2 structures of each 52 IMP variants, structurally aligned. **C:** lDDT scores for each residue after structural alignment with FoldMason, indicating level of structural conservation. **D:** Amino acid conservation score, as calculated using residue similarity and the bio3d sub matrix. The Multiple Sequence Alignment is found at Figshare: 10.6084/m9.figshare.28440992.

### Non-human reservoirs of *bla*IMP carbapenemase genes

We then adopted a One Health approach by analysing sources of *bla*_IMP_ genomes to determine the potential contribution of environmental or animal reservoirs to *bla*_IMP_ dissemination. Genomes of human origin accounted for the majority (4,051/4,556, 88.9%), while genomes of environmental and animal origin accounted for 10.0% (454/4,556) and 1.1% (51/4,556), respectively (**Fig. 7A**). Nine samples were unable to be classified. Most environmental genomes (353/454, 77.8%) came from healthcare settings, with at least 128/353 (36.3%) coming from hospital aquatic environments (**Fig. 7B**, **Table S9**). Animal data were limited but *bla*_IMP-4_ and *bla*_IMP-38_ were detected in Australian seagulls (n=31 and n=1, respectively) (36), and *bla*_IMP-27_ in US and Canadian genomes of pig origin (n=4 and n=1, respectively). 65/1,381 (4.7%) IMP-clusters were found across multiple source categories (**Fig. 7A**), most notably *bla*_IMP-26_ *P. aeruginosa* ST235 and *bla*_IMP-4_ *S. marcescens*. Only three IMP-clusters had genomes of both human and animal origin (all birds), including *bla*_IMP-4_ *E. coli* ST58, *bla*_IMP-27_ *P. rettgeri* spp. 2 CG5 and *bla*_IMP-45_ *P. aeruginosa* ST313.

**Fig. 7:**
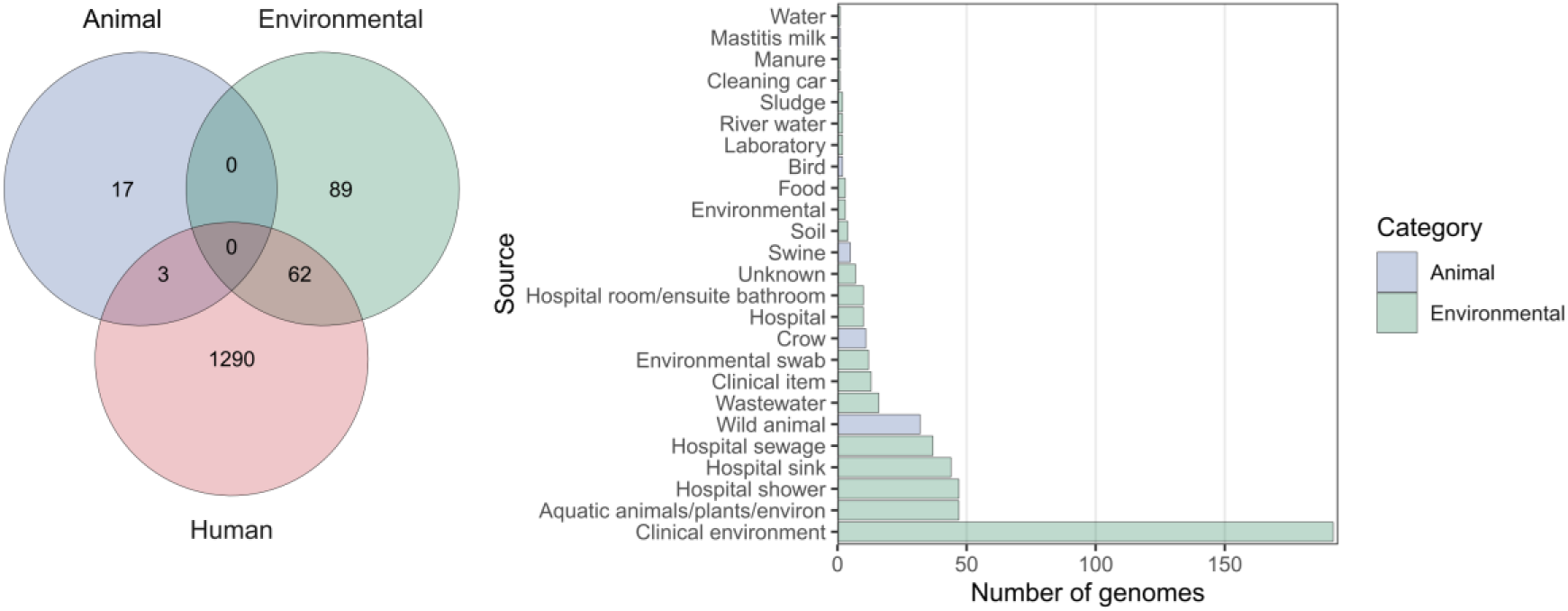
Human, animal and environmental origins of *bla*_IMP_ genomes. **A:** Venn diagram showing the intersections of IMP-clusters (as previously defined) between genomes of human, environmental and animal origins, indicating either clonal spread between categories or discrete isolation. **B:** Number of genomes isolated from specific non-human sources.

## Discussion

We have systematically determined the global distribution, diversity and epidemiology of 52 *bla*_IMP_ variants across 4,556 publicly-available genomes. This enabled us to reconstruct a complete picture of the emergence of IMP carbapenemases as a global and multi-faceted problem across all geographic regions. Previous studies have either focused on a single *bla*_IMP_ variant or geographical site (5,19,22,37–39), leaving gaps in our understanding of the genomic epidemiology of these genes. Our analysis has revealed the under-recognised diversity and global distribution of *bla*_IMP_ carbapenemase genes. We have shown that *bla*_IMP-1_ and *bla*_IMP-4_ were the most common variants due to their early emergence (40,41) and wide dissemination, achieving global endemicity along with *bla*_IMP-7_, *bla*_IMP-8_, *bla*_IMP-13_ (**Fig. 1**). In contrast, several *bla*_IMP_ variants displayed distinct regional patterns including *bla*_IMP-26_ (**Fig. 2A**) in Southeast Asia and *bla*_IMP-27_ in North America (**Fig. 2C**), achieving regional endemicity. These patterns have been previously unrecognised and challenges earlier perceptions that IMP carbapenemases are largely confined to the Asia-Pacific region (42).

Horizontal gene transfer, more than clonal spread, enabled global dissemination of *bla*_IMP_ genes. While previous work has demonstrated associations of *bla*_IMP_ and *bla*_GES-5_ genes with integrons (19,43,44) and co-location with IS (45,46), we demonstrate these intersections systematically at scale. We found that *bla*_IMP_ genes were almost invariably embedded in class 1 integrons, frequently flanked by IS. This genetic context promoted *bla*_IMP_ mobility, enabling entry into a vast array of at least 52 plasmid clusters. These included broad host-range IncHI2A, IncC, and IncN plasmids, all found to carry multiple *bla*_IMP_ variants. The coupling of integrons and IS with these broad-host range plasmids likely drove interspecies spread of *bla*_IMP_ genes into diverse bacterial hosts, specifically *Enterobacterales* (e.g. *Enterobacter* spp., *Klebsiella* spp. but also *Providencia* spp. and *Proteus* spp.). The regional *bla*_IMP-27_ gene was not associated with any known plasmids using our approach (which relied on completed plasmid references), but has been shown to be associated with the conjugative pPM187 (IncX8 replicon type) and pPR1 (no known replicon type) plasmids, allowing experimentally-verified inter-species spread (47).

Despite horizontal gene transfer being a key driver of *bla*_IMP_ spread, we also noted that proliferation of certain bacterial host lineages was an important contributor to *bla*_IMP_ dissemination. *bla*_IMP-1_ *P. aeruginosa* ST235 was a successful global lineage but other lineages were more geographically limited. Instead, we noted numerous local outbreaks and limited inter-country spread of successful plasmid-lineage combinations that we previously termed ‘propagators’ (19). We identified at least 35 propagators that sustained and amplified local *bla*_IMP_ outbreaks (e.g. *bla*_IMP-4_ *S. marcescens*-IncC, *bla*_IMP-1_ *E. hormaechei*-IncHI2A). This is in contrast to other carbapenemases such as KPC and OXA-48, which are associated with globally disseminated clones such as *K. pneumoniae* ST258 and ST11, respectively (48). In addition, we propose the idea of ‘connectors’: lineages that can accept multiple plasmids and serve as bridges for *bla*_IMP_ gene transfer through mobile genetic elements, without themselves causing local outbreaks. We identified 64 such lineages such as *E. hormaechei* ST78 and *E. coli* ST131. The interplay of propagators and connectors helps explain how *bla*_IMP_ genes can disseminate and also repeatedly establish in new hosts. The *bla*_IMP_ threat is therefore multidimensional: it spreads by many local expansions rather than a single dominant lineage crossing borders, thus posing difficulties for both detection and control.

Beyond the various levels of *bla*_IMP_ gene transfer, we also analysed the structures of IMP enzymes to examine whether adaptive changes in the enzymes themselves may be contributing to spread. Despite the diversity of 52 known IMP variants, we detected convergent evolutionary patterns, with repeated missense mutations at specific sites. These changes likely offer functional advantages by altering carbapenem hydrolysis activity and thus minimum inhibitory concentrations (30,33–35). For example, IMP-26, which we identified as key regional endemic clone in Asia, is more effective at hydrolysing meropenem and doripenem than older variants and thus displays a broader and more effective carbapenemase phenotype (49). While we cannot determine which selective pressures led to the rise of each *bla*_IMP_ variant, we speculate that this could be driven by antimicrobial usage (30) and species-specific susceptibility (50). The fact that similar mutations have arisen independently in different *bla*_IMP_ variants implies ongoing adaptive evolution. Similar convergent mutation patterns have been shown in the *K. pneumoniae* extended-spectrum β-lactamase *bla*_SHV_ (51). These evolutionary patterns raise concern that IMP enzymes may continue to evolve, not only with increasing carbapenemase activity but also potentially evading even the newest β-lactamase inhibitors.

Finally, we adopted a One Health lens to help understand and hopefully control *bla*_IMP_ spread. Although most genomes came from human clinical isolates, *bla*_IMP_ genes were also found in hospital environments (e.g., surfaces, wastewater), as well as animal samples (e.g., in cats and birds). Our fine-grained IMP-cluster analysis allowed detection of movement between these One Health categories, predominantly between human clinical isolates and clinical environments, and to a lesser extent between environmental/animal sources. This illustrates how healthcare-associated resistance genes can spill over into animals or the environment, creating secondary reservoirs. These reservoirs may, in turn, seed new infections back into humans, forming a cycle that blurs traditional epidemiological boundaries. The presence of *bla*_IMP_ genes in *Serratia* spp.*, Pseudomonas* spp. and other environmental bacteria also raises the possibility of environmental persistence. *bla*_IMP_ genes therefore have access to multiple ecological niches (human, animal, and environmental) that constantly interact, further complicating control efforts. Furthermore, non-clinical isolation sources are likely under-sampled, thus underestimating the true extent of non-human *bla*_IMP_ reservoirs and underscoring the need for a One Health approach for surveillance and control.

Our study had ambitious reach, aiming at analysing the global genomic epidemiology of *bla*_IMP_ genes, but this was also the source of several limitations. Our analysis relied on publicly-available genomes that represented biases in whole genome sequencing activity and genomic surveillance. This led to overrepresentation of countries with genomic infrastructure, with corresponding under-representation of low- and middle-income countries that are likely disproportionately impacted by infections caused by IMP carbapenemases (52). On a technical level, we were reliant on the quality of genomic data submitted. Most data were derived from short-read sequencing, introducing uncertainty for determining the genetic contexts of *bla*_IMP_ genes. We tried to compensate for this by conducting a mapping-based analysis but acknowledge the inherent limitations of this approach. Similarly, metadata were frequently poorly curated, thus preventing discovery of additional non-clinical spread and limiting geographic/temporal inferences. Moreover, the dataset includes 52 of 96 known IMP variants, leaving rare variants uncharacterised.

In summary, we have demonstrated that the emergence of IMP carbapenemases has largely ‘flown under the radar’, despite the establishment of *bla*_IMP_ variants with global endemicity (IMP-1, IMP-4, IMP-7, IMP-8, IMP-13), as well as regional endemicity (IMP-26 and IMP-27). We have shown that this global spread was the result of a complex interplay of genomic drivers of dissemination (at the mobile genetic element, plasmid and bacterial host lineage levels), with horizontal gene transfer playing a more substantial role than global spread of specific lineages. We noted convergent evolution in IMP carbapenemase enzymes, suggesting adaptation under evolutionary pressures and posing an ongoing challenge with the possibility of adapting to and conferring resistance to new antimicrobial treatments. Finally, we noted the presence of One Health reservoirs of IMP carbapenemases, although detailed analysis was limited by the small number of non-clinical genomes. These findings provide a detailed atlas of IMP carbapenemases and their global spread that also carry implication for other carbapenemases and emerging forms of antimicrobial resistance. In addition to casting light on the extent IMP carbapenemases have silently become a global problem, we have provided a roadmap for future interventions to disrupt their future spread. Our work highlights the need for more robust and sophisticated surveillance approaches that address gaps in low- and middle-income countries, incorporate methodology to detect plasmid and mobile genetic element transmission and conduct more systematic sampling of One Health reservoirs. We have carried out a systematic global analysis of IMP carbapenemases using three decades of data but the critical threat they pose now requires a proactive, real-time and co-ordinated public health response.

## Methods

### Genome acquisition, species identification and genotyping

NCBI Pathogen Isolate Browser (53) was searched using the term "AMR_genotypes:blaIMP*" to retrieve assemblies containing all IMP variants. This initial search identified a total of 4,063 genomes from 517 BioProjects. Of these, 3,177 assemblies were downloaded from Pathogen Isolate Browser and 886 reads were obtained from Sequence Read Archive (SRA) and assembled with Unicycler v.0.4.8 (54) with standard parameters.

To identify additional publicly available assemblies, protein accession numbers for all IMP variants were obtained and queried against NCBI GenBank databases using BLAST v2.15 (55). Assemblies retrieved from this search were compared against the Pathogen Isolate Browser dataset, and non-duplicates were retained, resulting in an additional 583 assemblies. Collectively, this approach yielded a total of 4,556 assemblies, which were used for all subsequent analyses. Metadata was extracted from the BioSample accessions. In cases where no ‘collection date’ was available and a BioProject did not contribute ≥50 isolates, the date of sequence upload was used as a proxy. Assembly quality was checked using Quast v5.2.0 (56) and species identification was performed using Speciator (57). Genomes were annotated using Prokka v1.14.6 (58). We then performed resistance gene detection with AMRFinderPlus v3.12.8 (59). We determined *in silico* multi-locus sequence type (ST) using ‘mlst’ v.2.19.0 (60). All inconclusive ST calls were checked with SRST2 v0.2.0 (61).

We note that several key *bla*_IMP_-containing species such as *Providencia rettgeri*, *Providencia stuartii, Morganella morganii* and *Proteus mirabilis* do not have well-defined MLST schema. As such, we employed a two-tiered approach to classify lineages for this study. Initially, untyped species were assigned MASH clusters if they were within ≤0.05 of each other to accurately group independent species (62), then PopPUNK v4.2.0 (63) was used on each of these MASH clusters to assign lineages for within-species comparisons. The ‘create-db’ function was used with the following options: ‘--sketch-size 1000000 --min-k 15 --max-k 29 --qc-filter prune’. Then the ‘fit-model’ function was used with the following options: ‘bgmm --ranks 1,2,3,5 --graph-weights’. Various --K values were used to obtain the best-scoring model fitting. Additionally, the ‘poppunk_visualise’ function was used, with the ‘--distances’ and ‘--previous-clustering’ to output a neighbour-joining core tree.

We identified that species identified as *Providencia rettgeri* and *Providencia stuartii* were considerably divergent and the taxonomy of these species requires detailed revision, with five and two independent species groupings outside the 0.05 standard threshold. These displayed MASH distances of up to 0.19 ‘within species’, indicating that although these ‘species’ are related, they are distinct species or potentially genera. However, as this is beyond the scope of our current work, we refer to these as *Providencia rettgeri* spp. 1, *Providencia rettgeri* spp. 2 etc.

For assemblies without recorded sequencing technology, a heuristic approach was adopted: assemblies with fewer than 20 contigs were classified as long-read or hybrid, while those with 20 or more contigs were marked as short-read. We assumed that long-read sequencing or assemblies with both short- and long-read data typically yields fewer contigs compared to short-read sequencing data alone. The United Nations geoscheme was used to classify countries into regions and subregions. This resulted in a dataset comprising 536 long and 4,022 short read assemblies.

### Assembly dereplication and IMP-cluster definition

To determine the impact of potential clonal outbreaks, a dereplicated dataset was employed. Initially, all genomes were split into their respect STs/CGs, then pairwise SNV distances were calculated using the ‘fasta’ command from SKA v1.0.0 (64), followed by the ‘distance’ command. Clonally-linked genomes were detected if two genomes had ≤5 pairwise SNVs per Mb. igraph v2.0.3 (65) was then used to perform graph-based clustering using pairwise SNV differences as edges, then membership data extracted for any isolates forming a distinct sub-clusters, allowing identification of putative spread clusters. ‘IMP-clusters’ were defined as genomes which shared the same *bla*_IMP_ variant, species, ST/CG, and were within the SNV threshold as previously defined.

### Plasmid analysis

Initially, the mob_recon function from MOB-suite v3.1.8 (25) was used to assign contigs to plasmids and identify replicon types and other information to long read assemblies only. These 433 plasmid-positive contigs were used to construct a custom *bla*_IMP_-positive plasmid database via the “mob_typer --multi", then “mob_cluster --mode build” commands. Short-read assemblies were then screened against this custom database with mob_recon.

### Genetic context of mobile genetic elements

Flanker v0.1.5 (66) was used to determine genetic context of *bla*_IMP_ genes with the following commands: “--flank both --window 10000 --gene blaIMP --include_gene --cluster”. For analysis of mobile elements, only sliced contigs with ≥6kb were used. Integrons were identified using Integron_finder v2.0.5 (67) with the “--local-max --func-annot --union-integrases” options. IS were screened using ISEScan v1.7.2.3 (68). ICEs were identified using the ICEberg 2.0 (69) database as a query against each genome with minimap2 v2.26 (70).

### Protein and structural analysis

SignalP v6.0 (71) was used to process the signal sequences from IMP protein sequences, then Clustal Omega v1.2.4 (72) was used to align these sequences. This was used as input to calculate amino acid conservation scores using the conserve() command from bio3d v2.4-4 (73), with the following parameters: ‘method="similarity", sub.matrix="bio3d"’. Colabfold v1.5.5 (74) was used to generate AlphaFold2 (75) structure predictions of mature IMP sequences, with the rank 1 structures kept. All structures were used as input for FoldMason v1.763a428 (76) to generate per-residue structural conservation lDDT scores. Scores were used to colour structures by residue using ChimeraX v1.8 (77) and custom scripts at https://github.com/bananabenana/ChimeraX_scripts (78).

## Statistical analysis

Statistics were performed in R v4.4.1 (79) and RStudio v2024.09.0 (80). The following R packages were used: tidyverse v2.0.0 (81), colorspace v2.1-1 (82), viridis v0.6.5 (83), ggh4x v0.2.8 (84), ggstream v0.1.0 (85), maps v3.4.2 (86), scatterpie 0.2.4 (87), sf v 1.0-17 (88), rnaturalearth v1.0.1 (89), ggnewscale v0.5.0 (90), treemapify v2.5.6 (91), patchwork v1.3.0 (92), igraph v2.0.3 (65), qgraph v1.9.8 (93), ggraph v2.2.1 (94), ggforce v0.5.0 (95), ggalluvial v0.12.5 (96), ggtree v3.12.0 (97), treeio v1.28.0 (98) and aplot v0.2.3 (99). All R code can be found at Figshare: 10.6084/m9.figshare.28440992.

## Data availability

All data used in this study is available as supplemental material (Figs. S1-4, Table S1-9). Additionally, all analysis code is available at Figshare: 10.6084/m9.figshare.28440992, along with additional supplemental material. Custom scripts can also be found at https://github.com/bananabenana/residue_structure_colour_scripts.

## Author contributions

Conceptualization: NM, AYP

Data Curation: BRM, BV

Formal Analysis: BRM, BV, HAN

Funding Acquisition: NM, AYP

Methodology: BV, BRM, NM

Project Administration: NM, AYP

Resources: NM, AYP

Supervision: NM, AYP

Writing – Original Draft Preparation: BRM, BV, NM

Writing – Review & Editing: BRM, BV, HAN, AG-S, AYP, NM

Both first-authors (BV, BRM) can present as first-author on paper for grants and resumes.

## Acknowledgements

This research/work was supported by Monash eResearch capabilities, including M3 and Research Data Storage.

