## Supplemental material for "The rise and global spread of IMP carbapenemases (1996–2023): a genomic epidemiology study"

### Supplemental Figures

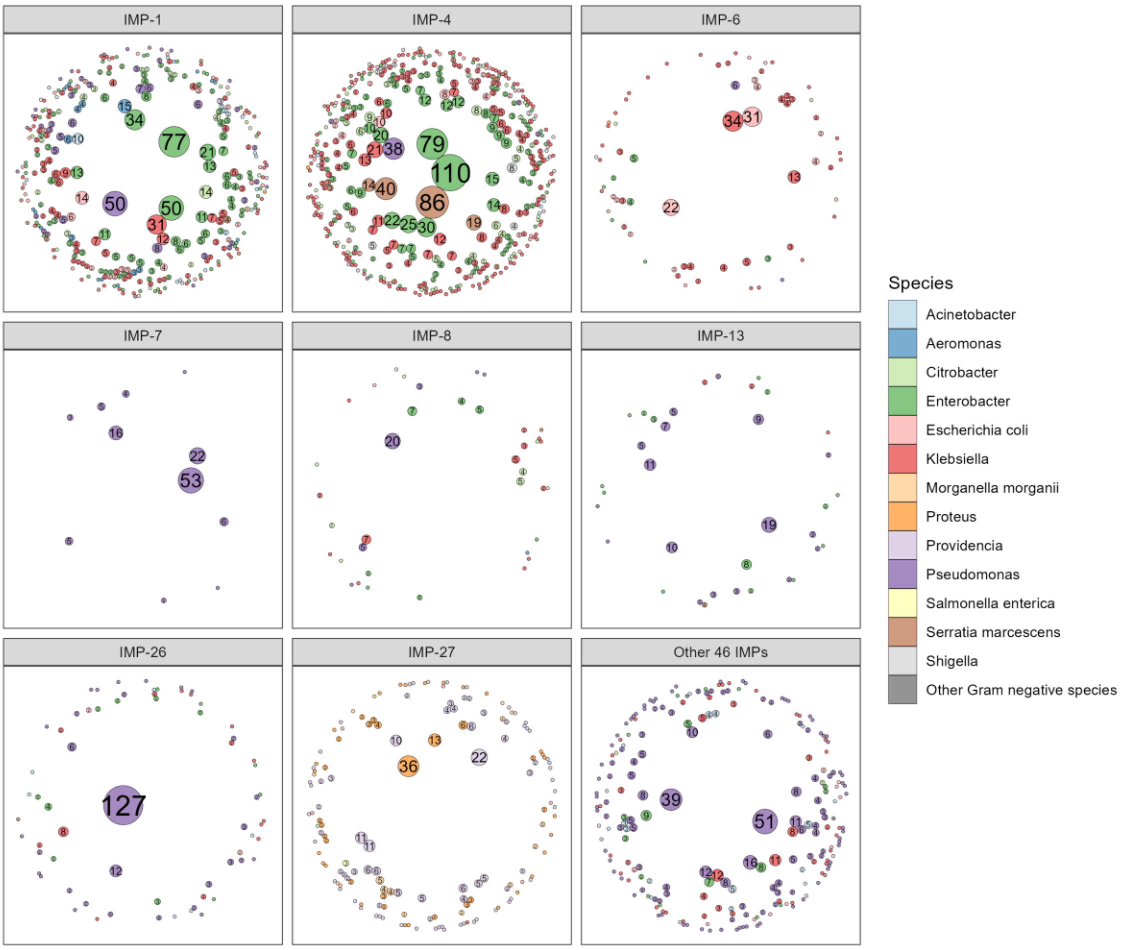


**Fig. S1: Size of IMP-clusters within dataset.** Number of genomes within IMP-cluster shown by size of bubble and text. No text is shown for IMP-clusters with n=1 genome.

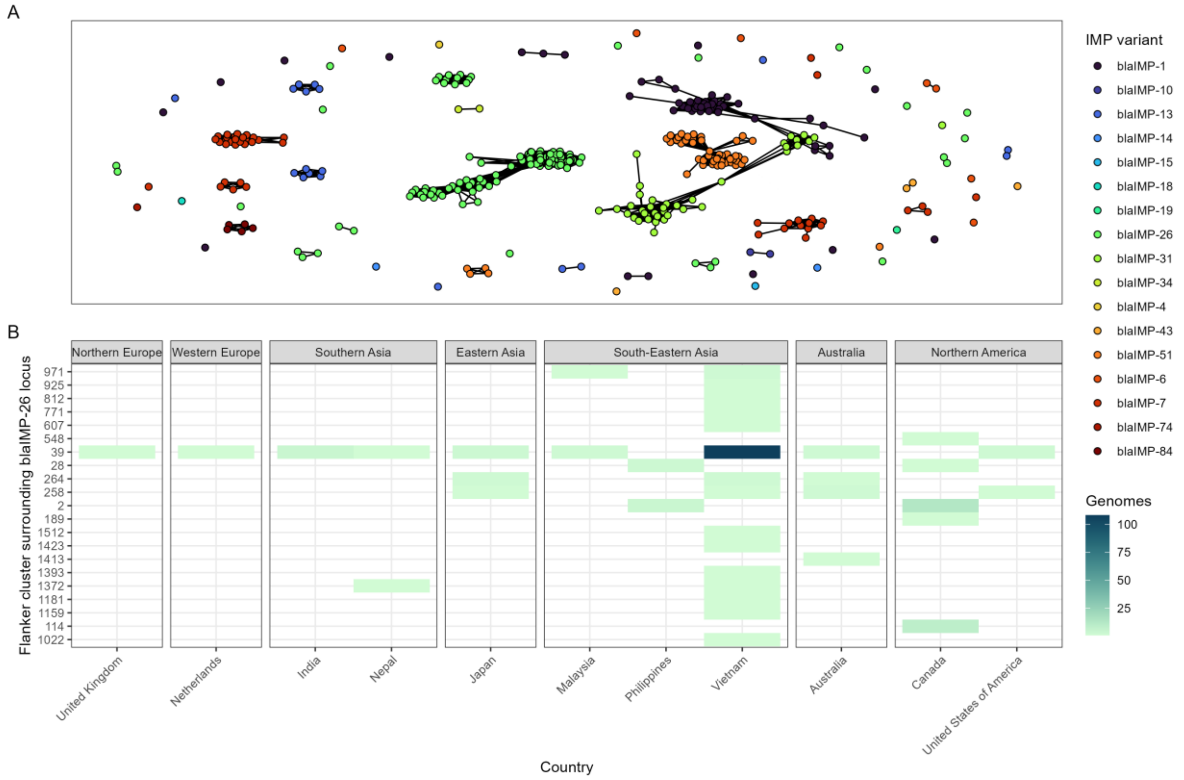


**Fig. S2: *bla*$_{IMP}$-carrying *P. aeruginosa* ST235 has global and diverse spread**

**A:** Network of clonally-linked *P. aeruginosa* ST235 genomes, coloured by *bla*$_{IMP}$ variant. **B:** Heatmap of IMP-26-containing *P. aeruginosa* ST235 showing regional-specific independent acquisition of the IMP-26 gene.

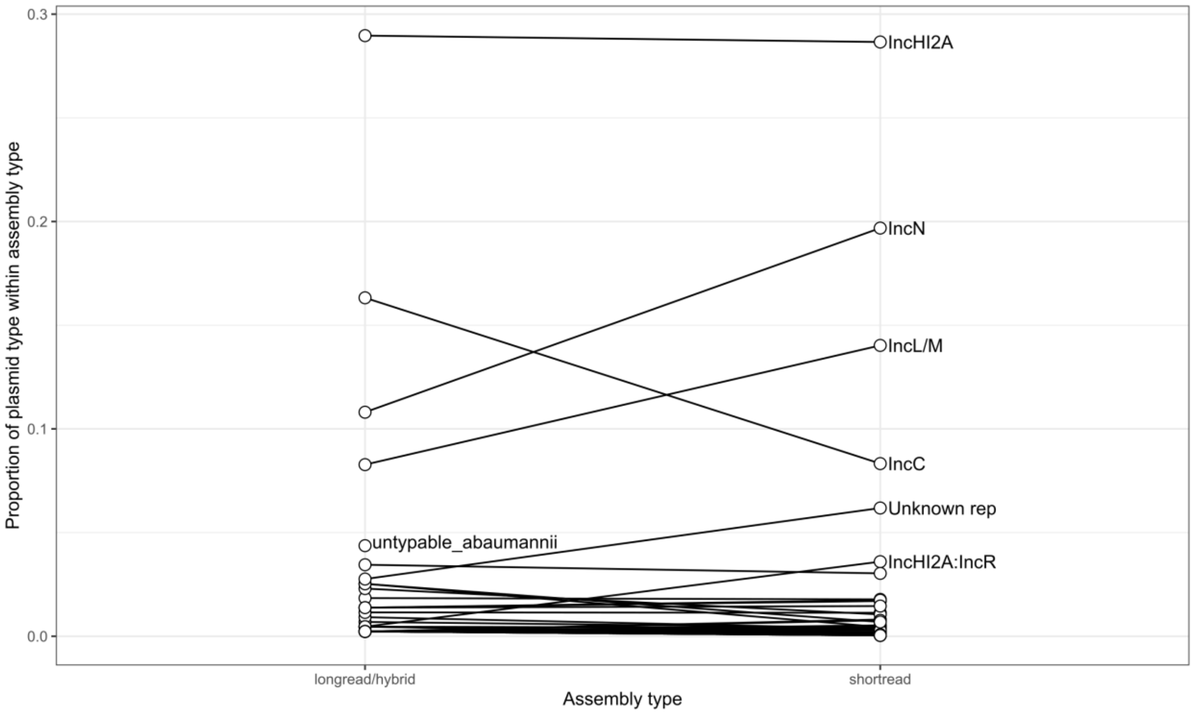


**Fig. S3: Proportion of plasmid clusters in long-read/hybrid dataset vs full dataset including short read draft assemblies.**

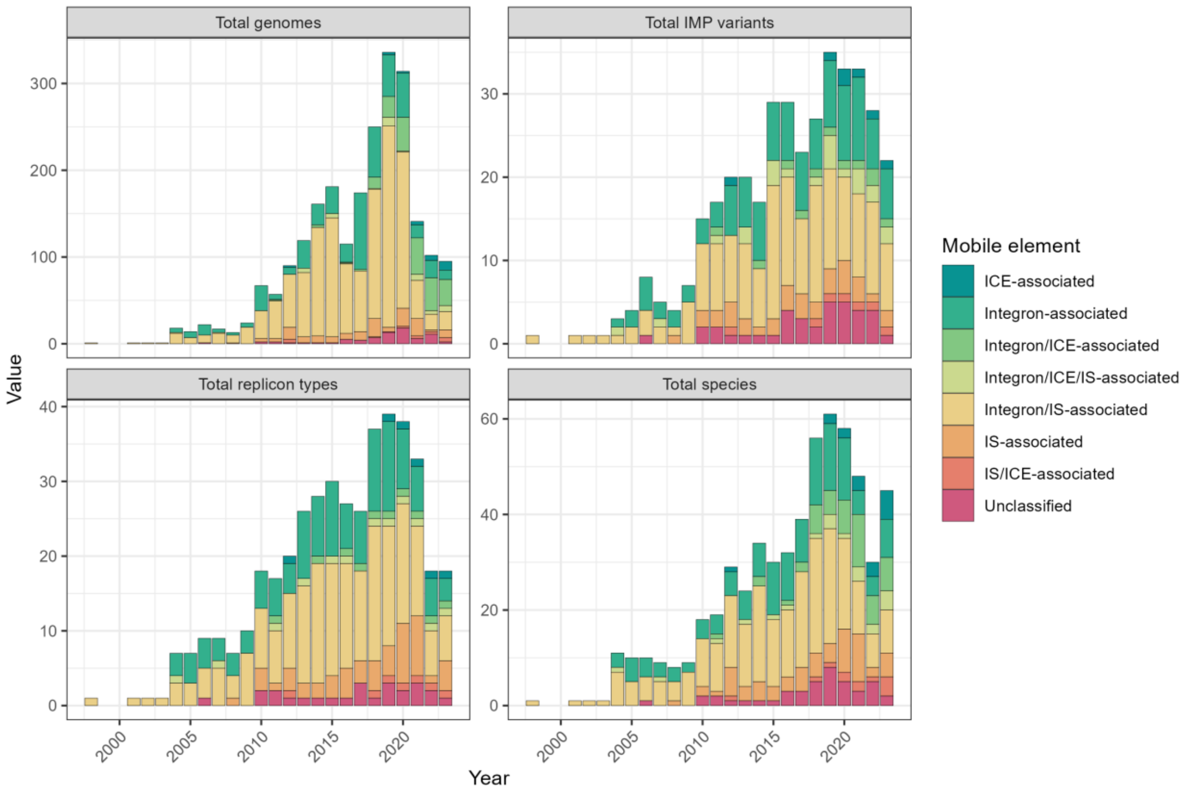


**Fig. S4: Mobile elements have been associated with IMP carbapenemases over time** Cumulative column graph showing the breakdown of associated mobile elements and their spread across numbers of species, *bla*$_{IMP}$ variants, total genomes and plasmid types. Raw data found in Table S1.

### Supplemental Table legends

**Table S1:** Table of all data used in this study, including accession numbers, analysis results and metadata.

**Table S2:** *bla*$_{IMP}$ variants over time, supporting information for Fig. 1.

**Table S3:** *bla*$_{IMP-26}$ and *bla*$_{IMP-27}$ supporting information for Fig. 3.

**Table S4:** *bla*$_{IMP}$ variants and their global distributions

**Table S5:** Association between bacterial lineages and *bla*$_{IMP}$ variants

**Table S6:** IMP-clusters and their global distribution and makeup

**Table S7:** Plasmid clusters and their spread across multiple countries and geographical regions for long read only genomes

**Table S8:** 'Propagator' strain-plasmid pairings and 'connector' strains.

**Table S9:** Summary of isolation sources and genome counts, supporting information for Fig. 7
